# Performance of the Cue COVID-19 Molecular Test for Point of Care: Insights from a multi-site clinic service model

**DOI:** 10.1101/2022.08.12.22278567

**Authors:** Anu Rebbapragada, Lane Cariazo, David Elchuk, Hossam Abdelrahman, Dang Pham, Nirochile Joseph, Elena Gouzenkova, Harpreet Gill, Peter Blecher

## Abstract

The COVID-19 pandemic highlighted the critical need for rapid and accurate molecular diagnostic testing. The Cue COVID-19 Point of Care Test (Cue POCT) is a nucleic acid amplification test (NAAT), authorized by Health Canada and FDA as a POCT for SARS-CoV-2 detection. Cue POCT was deployed at a network of clinics in Ontario, Canada with n=13,848 patrons tested between July 17, 2021 to January 31, 2022. The clinical performance and operational experience with Cue POCT was examined for this testing population composed mostly of asymptomatic individuals (93.7%). A head-to-head prospective clinical verification was performed between July 17 to October 4 for all POCT service clients (n= 3037) with paired COVID-19 testing by Cue and RT-PCR. Prospective verification demonstrated a clinical sensitivity of 100% and clinical specificity of 99.4% for Cue COVID-19 POCT. The lack of false negatives and low false positive rate (0.64%), underscores the high accuracy (99.4%) of Cue POCT to provide rapid PCR quality results. Low error rates (cancellation rate of 0% and invalid rate of 0.63%) with the current software version were additionally noted. Together these findings highlight the value of accurate molecular COVID-19 POCT in a distributed service delivery model to rapidly detect cases in the community with the potential to curb transmission in high exposure settings (i.e. in-flight, congregate workplace and social events). The insights gleaned from this operational implementation are readily transferable to future POCT diagnostic services.

## Introduction

Severe acute respiratory syndrome coronavirus (SARS-CoV-2) emerged as a pandemic threat in 2020 and continues to pose a global challenge with sustained waves of global infection, hospitalizations, and deaths due to respiratory infections. **(1)**. Due to the severity and heightened contagiousness of different SARS-CoV-2 variants, rapid and accurate testing is urgently required to reduce transmission & morbidity in high exposure settings (i.e. healthcare, in-flight airline cabinet, congregate workplaces and social gatherings) **(2,3,4,5)**. Detection of SARS-CoV-2 RNA by nucleic acid amplification tests (NAAT) such as Reverse transcription-polymerase chain reaction (RT-PCR) is the most common method **(4)**. However, lab-based RT-PCR is disadvantaged by transportation to a central testing site, longer analytical process (often 4-6 hours to perform specimen prep, extraction, detection) and result analysis. These inherent limitations with lab-based testing hamper the objectives of rapid case detection, enactment of infection control precautions, and early treatment. Point of Care Testing (POCT) holds the potential for rapid detection of COVID-19 including screening of asymptomatic populations and the initiation of informed actions in a cost-effective and clinically efficacious manner **(5,6,7,8,9)**.

The Cue COVID-19 molecular test (Cue POCT) is one of a handful of rapid molecular POCT that have received emergency use authorization (EUA) by US Food and Drug Administration (FDA) and Health Canada Emergency Interim Order (IO) to detect SARS-CoV-2 **(6,10,11)**. This report describes the performance of the Cue POCT at a network of clinics in Ontario, Canada providing COVID-19 screening services to asymptomatic individuals for the purposes of reassurance, travel permission, or entry into workplace and congregate events. The insights gleaned from test sensitivity, specificity and operational parameters can inform the implementation of future POCT diagnostic services.

## Methods

### Testing Centres

Specimen collection and Cue COVID-19 POCT were performed at a network of FH Health Clinics in Ontario Canada by registered nurses and RT-PCR testing was conducted at the FH Health Laboratory, licensed by the Ontario Ministry of Health and accredited under ISO 15189 standards. Clinic sites included densely populated urban locations, as well as suburban settings. Ontario provided free COVID testing services to symptomatic individuals who met eligibility criteria for public services. Patrons booked a clinic appointment digitally (via web or app) and upon arrival at the clinic, FH Health administrators verified patron identity, and collected information on symptom and vaccination status **(Table 1)**. The data presented in this manuscript represent a prospective verification of test performance of the Cue COVID-19 Point of Care Testing system versus RT-PCR leveraging results gathered from provision of routine testing services. This study methodology was reviewed by Advarra Review Board and was considered to be exempted. They noted the framework of this comparison (where it was conducted, information provided to paying patrons, anonymized data analysis and report) fulfill the criteria for Exemption under the Common Rule (45 CFR subpart A).

**Table 1:** Age demographics of Cue COVID-19 Test Population. a) Parallel testing period from July 17 to October 4 2021, n=3037 Cue patrons with parallel Cue and RT-PCR results; b) Entire period from July 17 2021 to Jan 31, 2022, n= 13,849 Cue patrons

| Age demographic of patients July 17 - Oct 4 (n=3,037) |  |  |
| --- | --- | --- |
| Birth Year | Category | Number of Patients |
| 2018-2021 | Baby (0-3) | 57 |
| 2004-2017 | Youth (4-17) | 259 |
| 1982-2003 | Adults (18-39) | 1,197 |
| 1962-1981 | Middle Aged (40-59) | 1,090 |
| <1961 | Seniors (60+) | 434 |
|  | **Total** | **3,037** |

**b.**
| Age demographic of patients July 17 - Jan 31 (n=13,849) |  |  |
| --- | --- | --- |
| Birth Year | Category | Number of Patients |
| 2018-2021 | Baby (0-3) | 191 |
| 2004-2017 | Youth (4-17) | 1,027 |
| 1982-2003 | Adults (18-39) | 5,637 |
| 1962-1981 | Middle Aged (40-59) | 5,060 |
| <1961 | Seniors (60+) | 1,933 |
|  | **Total\*** | **13,848\*** |

n= All Patrons tested from July 17 2021 to Jan 31, 2022.
\*One patron did not indicate DOB, hence excluded from the age stratification. After October 4, only patrons with either positive, cancelled or invalid Cue results had an additional swab collected for lab-based RT-PCR.

### Cue COVID-19 Molecular Test at FH Health Clinics

The Cue COVID-19 POCT system was utilized across 7 FH Health clinics for patrons who booked the “Express” service with results Turn Around Time (TAT) of 1-6 hours. Cue COVID-19 POCT was performed as per the approved Instructions for Use (IFU, **11**) by collecting an anterior nares specimen with the Cue wand, insertion into a Cue cartridge readied for testing on a Cue Device and reviewing results for the device on the paired Cue Health app. Use of the Cue POCT requires approximately 25 minutes (3 minutes pre-analytical and 22 minutes analytical time).

### Laboratory RT-PCR Tests

After collection of the anterior nasal sample for Cue testing with the Cue wand, nurses collected a bilateral deep nasal specimen with an FDA EUA collection device (Bioer, Hangzhou Bioer Technology co., Ltd) using a sterile nylon fiber non-flocked swab and placed into a tube containing 2 mL molecular transport media with guanidine for transportation to the lab for RT-PCR testing. RT-PCR specimens were tested within 6-12 hours of collection on one of the following RT-PCR assays: (i) Osang GeneFinder COVID-19 (FDA EUA and Health Canada IO), (ii) ThermoFisher TaqPath™ COVID-19 Combo Kit (FDA EUA and Health Canada IO) or, New England Biolabs Luna One Step SARS-CoV-2 RT-qPCR Multiplex Assay kit. Turn Around Time (TAT) for FH Health’s lab-based RT-PCR testing offered results within 12 or 24 hours, as “same day” or “next day” services.

Three different assays were validated to provide flexibility and insulate against supply chain constraints. Assays were utilized based on current reagent stock levels and workflow. RT-PCR specimens were primarily tested with the FDA EUA Osang GeneFinder COVID-19 assay (Osang). When Osang reagents were not available, RT-PCR specimens were tested either with the FDA EUA ThermoFisher TaqPath™ COVID-19 Combo Kit (TaqPath) or the New England Biolabs Luna SARS-CoV-2 RT-qPCR Multiplex Assay kit (Luna).

Samples for all assays were tested on a 96-well PCR plate with three external quality control wells on each plate. Positive SARS CoV-2 detection was reported when 2 or more viral targets were detected with amplification Ct values under 37 for the TaqPath assay or Ct values under 40 for the Osang and Luna assays. Indeterminate SARS CoV-2 detection was reported when only 1 viral target was detected with Ct values under 37 for the TaqPath assay or Ct values under 40 for the Osang and Luna assays. The amplification curves for all positive and indeterminate samples were evaluated by certified medical lab technologists. Samples reported as negative fulfilled the criteria for internal process control detection (MS2 on TaqPath) or human RNaseP (GeneFinder and Luna assays) indicating proper specimen collection, operator setup and assay system performance when viral target genes were not detected. Samples with no RNaseP amplification were reported as Invalid to indicate an issue with specimen collection and to trigger recall of the patron for repeat collection and testing. RT-PCR assays were verified to detect 2 copies of SARS CoV-2 per reaction. The Osang GeneFinder and New England Biolabs Luna assays demonstrated enhanced LoD to reliably detect 1 copy of SARS CoV-2 per reaction.

### Result Reporting

All results were digitally provided directly to patrons as a PDF report via FH Health’s proprietary Clinic and Laboratory Information System. The accuracy, reliability, PHIPPA compliance and security of digital reporting were verified prior to deployment and met the requirements of ISO 15189 accreditation standards.

### Cue POCT Performance Verification prior to implementation

#### Analytical Sensitivity

As per the IFU analytical sensitivity or limit of detection (LoD) of the Cue POCT was established by testing known amounts of purified viral culture (diluted in negative clinical nasal specimens) and claims a LoD of 20 copies when the Cue sample wand is used for specimen collection and direct testing on the Cue device **(10,11)**. This claimed LoD was verified at with a contrived panel of positive specimens prepared with Microbix™ REDx FLOQ® swab-based positive controls for the wild type and five variant strains of SARS CoV2 including, B.1.1.7-Alpha, B.1.351-Beta, B.1.617-Delta, P.1-Gamma and B.1.1.529-Omicron. Microbix™ REDx FLOQ® controls contain the whole genome of each SARS CoV-2 strain (estimated 35,000 copies/swab) and include human fibroblasts to permit evaluation of the specimen adequacy target detection (human RNAseP) used in the Cue POCT. A dilution series was prepared in 1ml viral transport medium (Microbix™ DxTM) to verify the reportable range and LoD for detection of wild type and five SARS CoV2 variants.

A Cue sample wand was dipped into each dilution to saturate sample absorption (approximately 50ul absorbed per wand) and tested on the Cue device. Results verified the claimed LoD of 20 copies per Cue wand for viral target gene detection.

#### Accuracy and Precision

Since residual RT-PCR specimens collected in molecular transport media could not be used to perform the Cue POCT (due to the presence of guanidine, which interferes with the Cue test reagents), accuracy was verified by testing a contrived panel of 20 positive specimens (prepared with the Microbix™ REDx FLOQ® Positive Control Swab) and 20 negative specimens (collected with the Cue wand from 20 COVID-19 negative staff members). Cue POCT precision was verified by testing at least 3 positive and 3 negative samples in duplicate for inter-device and inter-operator reproducibility. The Cue POCT system (reagents and reader device) met the acceptance criteria for accuracy and precision with 100% concordant results.

## Results

### Clinical Performance Verification of Cue POCT with parallel RT-PCR

#### Specimen disposition

A total of 3,368 tests were performed on the Cue Point of Care Test (POCT) from July 17 to October 4, 2021. From this set, n=3037 patrons had evaluable Cue and parallel lab-based RT-PCR results and were included for comparative analysis (2955 SARS-CoV-2-negative and 82 SARS-CoV-2-positive specimens).

Among the 3,368 tests performed, n=331 tests were excluded from comparative analysis due to invalid or cancelled Cue POCT results and, lack of a parallel RT-PCR specimen. 151 tests had an initial invalid (n=116) or cancelled (n=35) result representing a combined error rate of 4.48%. Re-testing these invalid and cancelled specimens as per the IFU yielded 135 valid results. Of the 116 invalid Cue tests, 6 could not be resolved and were sent to the laboratory for PCR testing. 25 of the cancelled Cue tests produced a negative test upon retesting. The remaining 10 cancelled tests could not be resolved, so RT-PCR samples were sent to the laboratory to obtain a valid result. Parallel samples for RT-PCR testing could not be collected from 180 individuals, including 2 Cue-positive patients, either because the patient could not return to the clinic or refused a parallel sample collection.

The majority of patrons were Asymptomatic (n=2998/3037, 98.7%) with the highest proportion of concordant positive results (n=61/63, 96.8%) (**Table 3)**. Symptomatic patrons (n=39, 1.28%) represented a lower proportion amongst positive detections (n=2/63, 3.17%).

As of October 5 2021, Cue test verification passed technical review by accreditation. After this point, parallel testing with RT-PCR was only performed for Cue patrons who tested positive or experienced two consecutive invalid or cancelled results. During the period from October 5, 2021 to January 31, 2022, n=10,811 Cue tests were performed, bringing the total number of Cue COVID-19 results for the entire period July 17 2021 to January 31, 2022) to n=13,848.

The Cue POCT demonstrated 100% clinical sensitivity (n=63/63) and high clinical specificity (99.4%, n=2955/2974), **Table 2**. The Cue Test demonstrated 99.4% concordance with RT-PCR testing, which provides high Positive Predictive Value in higher prevalence scenarios (97% when 20% prevalence) and 100% Negative Predictive Value. Asymptomatic individuals were the predominant client segment for FH Health during the July 17 to October 4 2021 paired testing period (98.7% asymptomatic, **Table 3**) and for the entire period through January 31, 2022 (93.7% asymptomatic, **Table 4**).

**Table 2:** Clinical Performance Characteristics of Cue COVID-19 vs. Laboratory RT-PCR testing, prospective parallel testing period from July 17 to October 4, 2021, n = 3037.

| **Cue Result** | **RT-PCR Positive** | **RT-PCR Negative** | **Total** |
| --- | --- | --- | --- |
| **Cue Positive** | 63 | 19 | **82** |
| **Cue Negative** | 0 | 2955 | **2955** |
| **TOTAL** |  |  | **3037** |

| **Test Performance** | **Cue POCT vs. RT-PCR**<br>**(n=3037 parallel tests)** |
| --- | --- |
| Sensitivity (63/63) | **100%** |
| Specificity (2955/2974) | **99.4%** |
| False Positive Rate (19/2974) | **0.64%** |
| Accuracy (TP+TN/total samples tested)<br>63+2955/3037 | **99.4%** |
| False Negative Rate (0/63) | **0%** |
| Positive Predictive Value | 76.8% (if 2% prevalence)<br>97% (if 20% prevalence) |
| Negative Predictive Value | 100% |

**Table 3:** Cue COVID-19 Patrons by Symptom Status during the parallel testing period, July 17 to October 4, 2021, n=3037. a) Number and proportion of Cue COVID-19 patrons who are symptomatic and asymptomatic; b) COVID Diagnosis by Symptom Status; c) Symptom status in True Positives, Cue COVID-19 and RT-PCR Positive; d) Symptom status in False Positives, Cue COVID-19 Positive and RT-PCR Negative

| Parallel Testing Period: July 17 - October 4, 2021 (n=3,037) |  |  |
| --- | --- | --- |
| Symptomatic status | Number of Patrons | Proportion |
| Symptomatic | 39 | 1.28% |
| Asymptomatic | 2,998 | 98.72% |
| Total | 3,037 |  |

**b.**
| Symptomatic |  |
| --- | --- |
| Negative | 37 |
| Positive | 2 |

| Asymptomatic |  |
| --- | --- |
| Negative | 2918 |
| Positive | 61 |
| False Positive | 19 |

| Symptomatic status | Number of Patrons |
| --- | --- |
| Symptomatic | 39 |
| Asymptomatic | 2,998 |

| c. |  |  | d. |  |  |
| --- | --- | --- | --- | --- | --- |
| Symptom Status | Cue and RT-PCR Positive | Rate | Symptom Status | Cue Positive and RT-PCR Negative | Rate |
| Asymptomatic | 61 | 96.8% | Asymptomatic | 19 | 100% |
| Symptomatic | 2 | 3.17% | Symptomatic | 0 | 0% |
| TOTAL | 63 |  | TOTAL | 19 |  |

**c) True Positives:** Asymptomatic Cue COVID-19 positive: n= 61, 96.8% Symptomatic Cue COVID-19 positive: n= 2, 3.17% The majority of positive patrons were Asymptomatic at our clinics. The high concordance between Cue COVID-19 and RT-PCR reflects high sensitivity of this technology.
**d) False Positives:** Asymptomatic Cue COVID-19 positive: n= 19, 100% Symptomatic Cue COVID-19 positive: n= 0, 0% All Cue COVID-19 False Positives were detected in Asymptomatic individuals, suggesting aberrant signal due to random primer interaction rather than cross-reactive detection with another respiratory pathogen.

**Table 4:** Symptom Status for Cue COVID-19 Patrons during the entire period July 17 2021 to January 31, 2022, n=13849. a) Number and proportion of Cue COVID-19 patrons who are symptomatic and asymptomatic; b) COVID Diagnosis by Symptom Status

| Symptomatic status July 17 - Jan 31 (n=13,849) |  |  |
| --- | --- | --- |
| Symptomatic status | Number of Patients | Proportion |
| Symptomatic | 863 | 6.23% |
| Asymptomatic | 12,986 | 93.77% |
| **Total** | **13,849** |  |

| Symptom Status | Positive | Negative* | Cue Positive<br>PCR Discordant |
| --- | --- | --- | --- |
| Symptomatic | 140 | 0 | 2 |
| Asymptomatic | 699 | 1 | 27 |
| **Total** | **839** | **1** | **29** |

For Cue Positive detections, the range of RT-PCR cycle threshold (Ct) values indicate a broad reportable range for samples that underwent parallel PCR testing during the July to October 2021 period (**Figure 1)** and the entire period through January 31, 2022 **(Figure 2**). RT-PCR Ct values for viral genes ranged from 13 to high 30s, demonstrating that the Cue POCT is able to detect virus throughout the infection life cycle i.e. from early symptom onset (post-exposure, early infection phase), peak viremia (high viral replication phase) and declining titres of viral load (recovery phase). Of note, 5 samples that tested positive by Cue had high Ct values (mean 34.5 to 38.3) with only a single gene detected by RT-PCR. These samples were categorized as “indeterminate” as per the laboratory reporting algorithm and recalled for follow-up testing. 1 patron did not return for testing. Two patrons tested positive when RT-PCR was performed on a new specimen collected a day after the original Cue positive result and two patrons had a recent history of prior infection indicating residual viral shedding during convalescence. The results suggest that the Cue POCT is able to detect SARS CoV2 in individuals with lower viral load, either at the early stages of a new infection or during the convalescent stage after symptoms have cleared.

**Figure 1:**
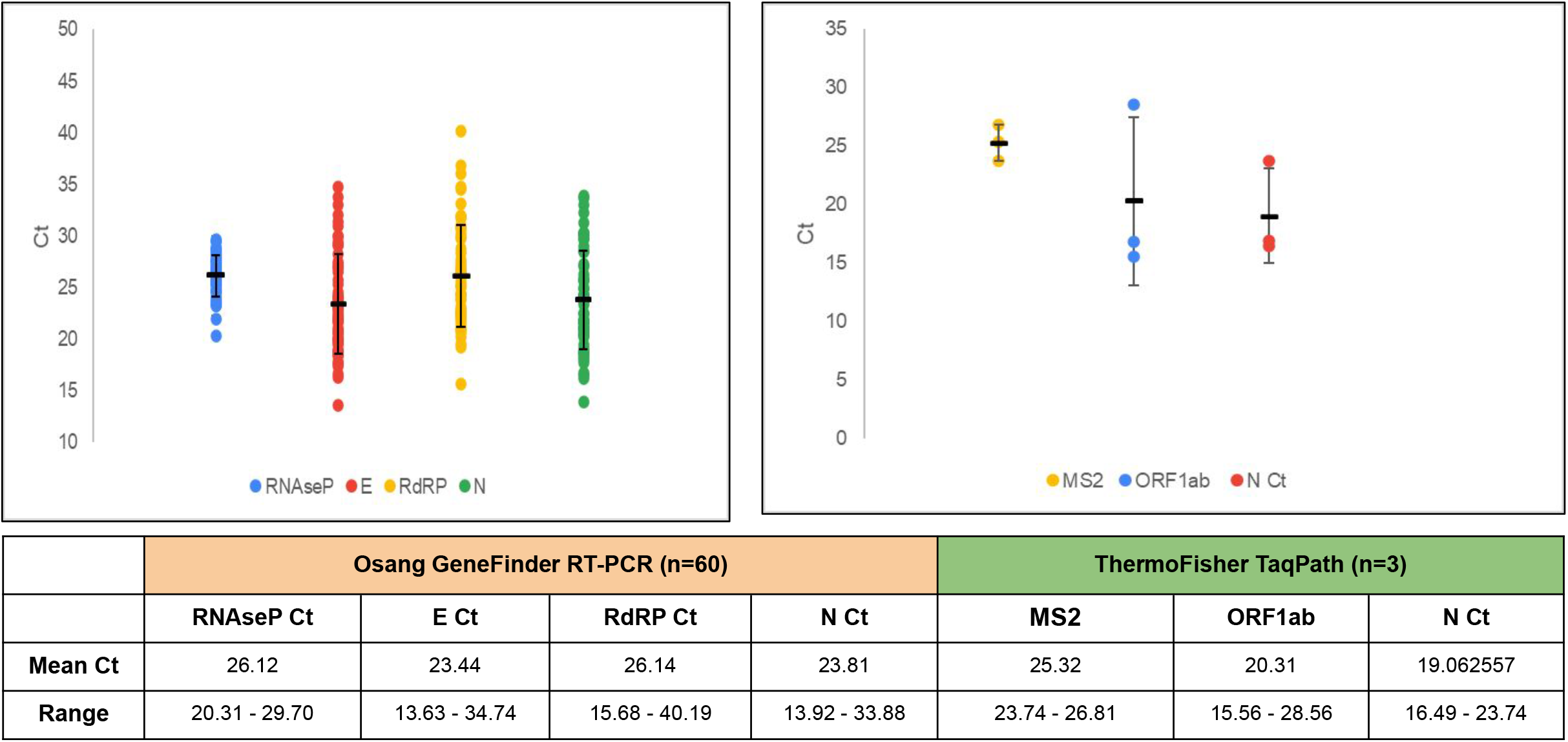
Cycle threshold (Ct) distribution for Cue True Positives, n= 63 from parallel testing period July 17 to October 4, 2021. a) Osang, n=60; Human RNAseP, Viral targets: E, RdRP, N; b) ThermoFisher TaqPath, n=3; MS2, Viral targets: ORF1ab, N. N.B. The S-gene was not detected in any of the samples tested by ThemoFisher. S-gene drop out represents Delta and Omicron variants

**Figure 2a:**
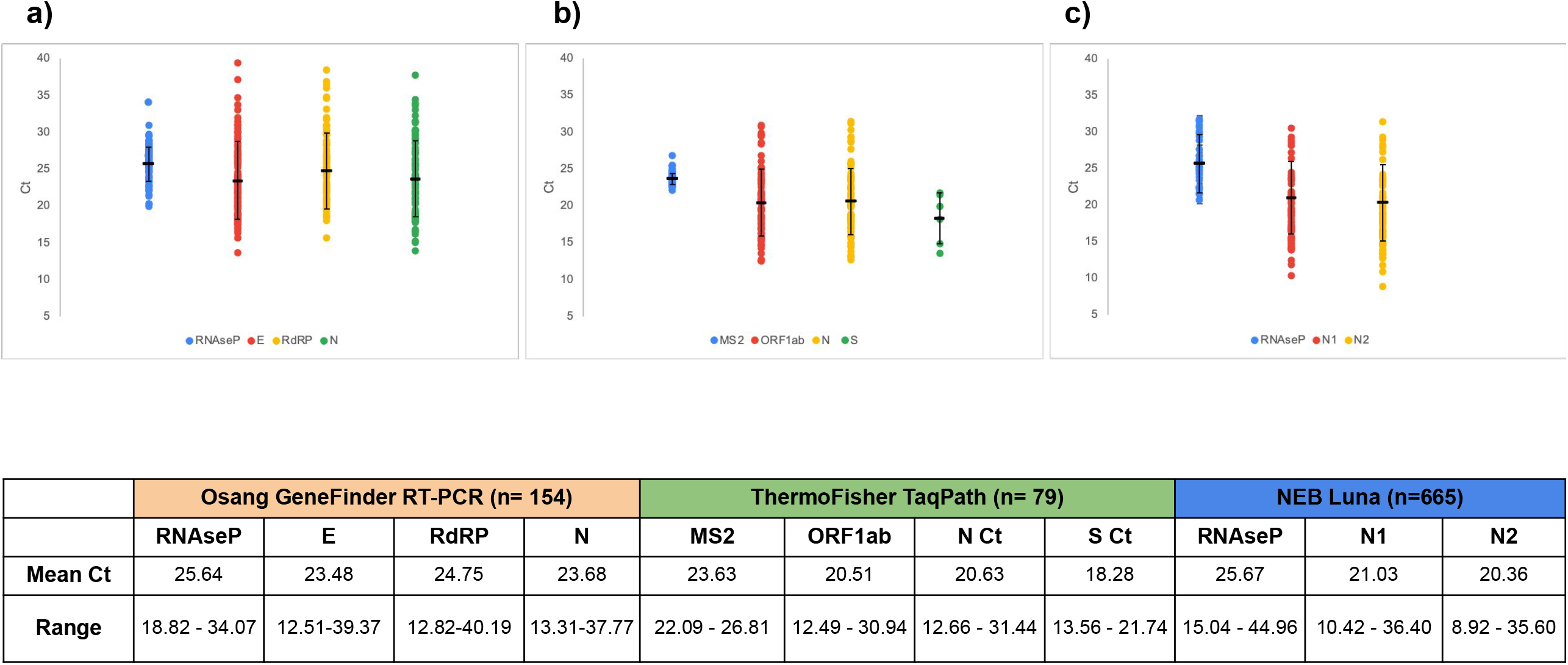
Cycle threshold (Ct) distribution for Cue True Positives, n=898 from July 17 2021 to Jan 31, 2022. a) Osang, n=154 Human RNAseP, Viral targets: E, RdRP, N; b) ThermoFisher, n=79: MS2, Viral targets: ORF1ab, N; c) Luna, n=665 Human RNAseP, N1, N2

**Figure 2b:**
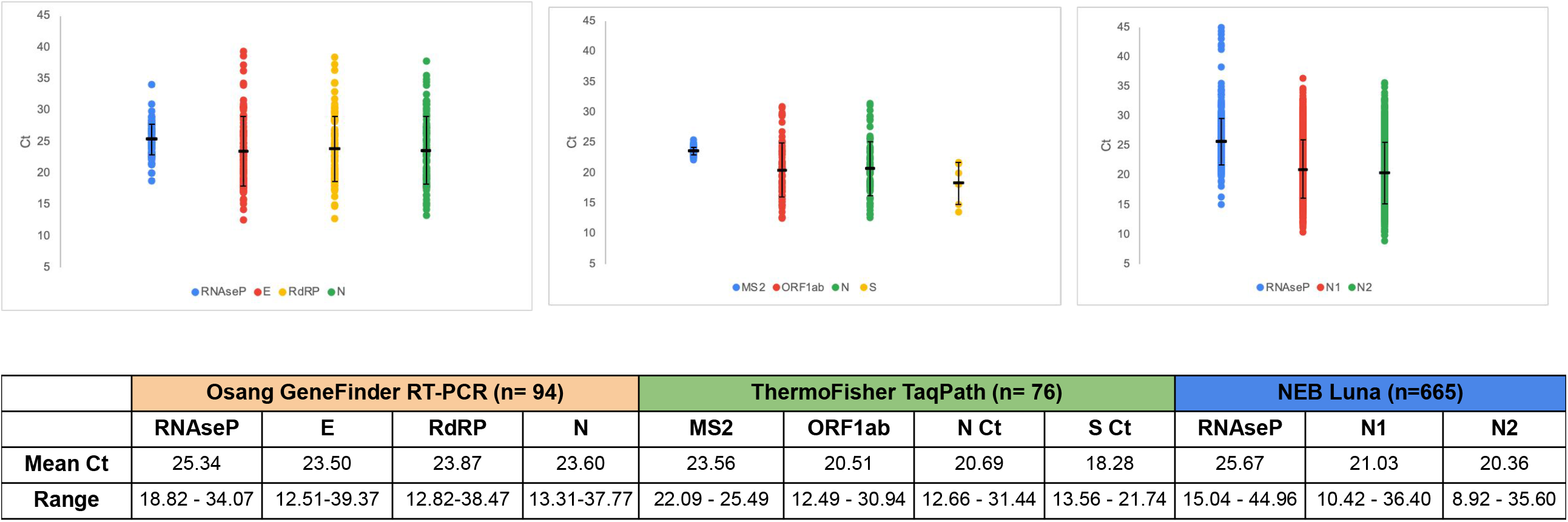
Cycle threshold (Ct) distribution for Cue True Positives, n=835 from October 5 2021 to January 31, 2022. a) n=**94** tested on Osang; order RNAseP, E, RdRP, N; b) n=**76** tested on TF; order MS2, ORF1ab, N. c)n=**665** tested on Luna, order RNAseP, N1, N2

The parallel testing data reveal a false positive rate of 0.64% (n=19/2974 results) for the period of July 17 to October 4 2021. All samples with discordant Cue POCT and RT-PCR results (i.e. Cue positive but RT-PCR negative) were re-tested by RT-PCR to confirm the validity of the negative RT-PCR result. All Cue POCT false positive results had valid human RNAseP detection by RT-PCR (**Figure 3**) and the three Quality Controls wells included on each PCR plate (extraction control, positive control and negative control) had valid/pass results indicating proper performance of RT-PCR reagents, equipment and operators. As of October 5 2021, parallel RT-PCR testing was only performed for patrons who tested positive on Cue COVID-19. The discordant rate between Cue positive and RT-PCR results for the entire period from July 17 2021 to January 31, 2022 was 0.21% (n=29/13849) with the majority of discordants identified in asymptomatic individuals across all testing periods (n=27/29) (**Table 3 and 4)**. These findings suggest that interfering substances in the nasal specimen or aberrant primer-dimer formations may have led to erroneous detection rather than cross-reactivity with another respiratory pathogen.

**Figure 3.**
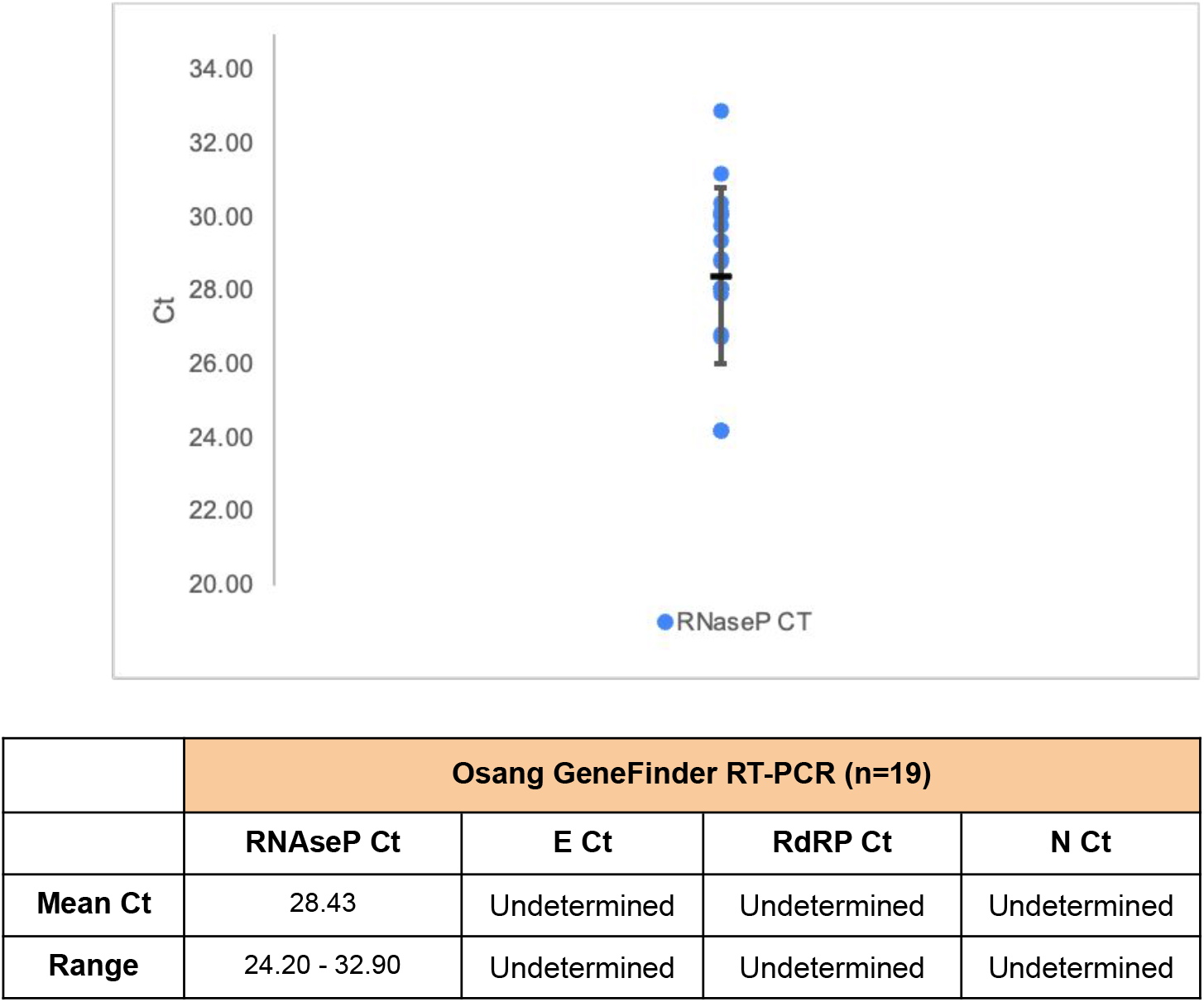
Cycle threshold (Ct) distribution for Human RNaseP detection in Cue False Positives, n=19 from parallel testing period July 17 to October 4, 2021.

#### Operational Metrics

Errors due to test cancellation or invalid results were significantly reduced with successive rounds of software upgrades and implementation of software upgrades (**Tables 5 and 6)**.

**Table 5:** Summary of Cue COVID-19 Result Categories. a) Parallel testing period from July 17 to Oct 4 2021, n=3037; b) Parallel testing period summary of only Valid Results, excluding Cancelled and Invalid categories; c) All results from July 17 2021 to Jan 31, 2022, n = 14331

| Summary of Cue Results July 17 - October 4 (n = 3,368) |  |  |
| --- | --- | --- |
| Result | Number of Tests | Rate |
| Negative | 3,133 | 93.24% |
| Positive | 84 | 2.49% |
| Invalid; Sample or Operator Error | 116 | 3.44% |
| Cancelled; Device Error | 35 | 1.04% |
| Cue Health Reporting App Error | 0 | 0% |
| **Total** | **3,368** |  |

**c.**
| Summary of Cue Results July 17, - January 31, 2022 (n=14,331) |  |  |
| --- | --- | --- |
| Result | Number of Tests | Rate |
| Negative | 12,585 | 87.81% |
| Positive | 1,154 | 7.95% |
| Invalid; Sample or Operator Error | 435 | 3.11% |
| Cancelled; Device Error | 157 | 1.13% |
| Cue Health Reporting App Error | 0 | 0% |
| **Total** | **14,331** |  |

**b.** Summary of results July 17 - October 4, 2021 (valid tests only)
| Categories | Number of patrons |  |
| --- | --- | --- |
| Negative | 2,955 | 97.30% |
| Positive | 82 | 2.70% |
| **Total** | **3,037** |  |

**Table 6:**
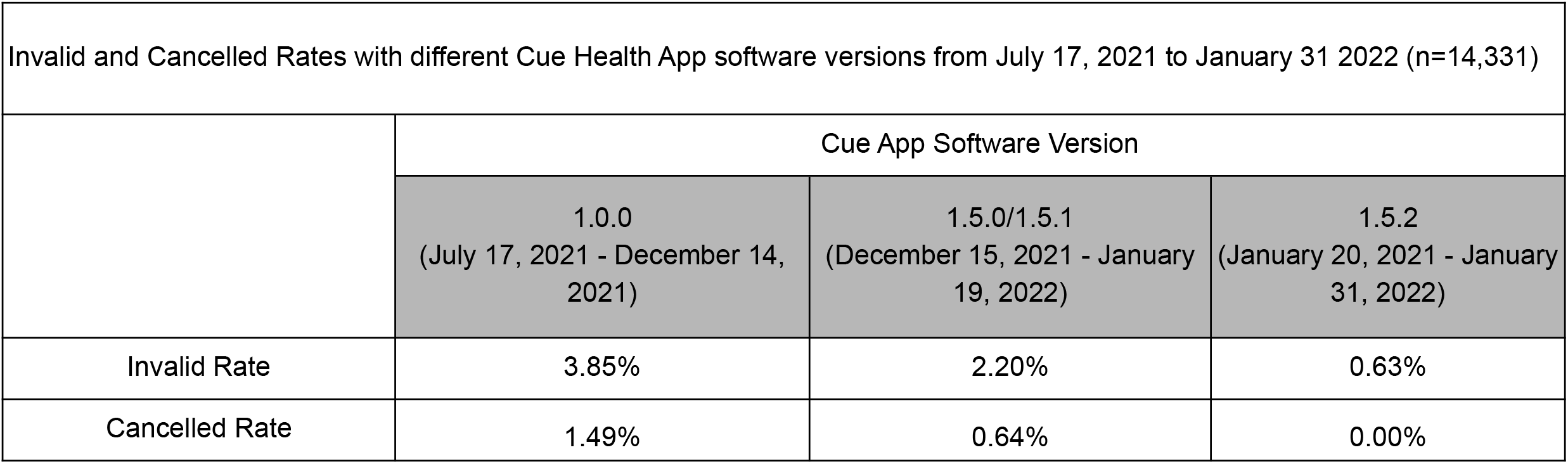
Cue COVID-19 Invalid and Cancelled Rates with different software versions from July 17 2021 to January 31, 2022.

The Cue Health App ran firmware updates starting in mid-December 2021 which reduced the occurrence of false positives by improving detection of mechanical issues related to wash buffer flow failure from the original version 1.0.0 used in July 2021; instead of reporting a false positive result, the safety update detects failure of this important process step and cancels the test. Upon implementation of serial firmware updates the false positive rate decreased further, from 0.49% to 0.39% (v.1.0.0 in December to January 19, 2022, v.1.5.0) and to 0.15% after January 20, 2022 (v.1.5.2).

Invalid errors are mainly due to improper specimen collection (inadequate sample obtained), operator errors in process or cartridge chemistry issues which can potentially compromise human RNaseP amplification. Invalid rates decreased from 3.85% to 2.2% to 0.63% corresponding to upgrades from versions 1.0.0 to 1.5.0 to 1.5.2, respectively.

Cancellation errors are due to safety check error (test cancels early upon initiation of process), component error or lack of detection of flow over sensor after lysis & amplification.

The cancellation error rate fell from 1.49% to 0.6% to 0% with serial software upgrades from versions 1.0.0 to 1.5.0 to 1.5.2, respectively.

Fewer than 1% of the study participants had test results delayed by over 2 hours due to test cancellation or repeated invalid results, thus minimizing the necessity to collect a new specimen or direct samples to the lab for RT-PCR testing (**Table 7**).

**Table 7:** Cue Results Turn Around Time, number and proportion. a) Parallel testing period from July 17 to October 4, 2021, n=3037; b) Entire period from July 17 2021 to January 31, 2022, n=13849

| Summary of TAT july 17 - oct 4 (valid tests only) n=3,037 |  |  |
| --- | --- | --- |
| Cue TAT | Number of Patrons | Rate |
| <30 min | 2,931 | 96.51% |
| <60 min | 101 | 3.33% |
| >2hr | 5 | 0.16% |
| Total | **3,037** |  |

| Summary of TAT july 17 - jan 31 (all patrons) n=13,849 |  |  |
| --- | --- | --- |
| Cue TAT | Number of Patrons | Rate |
| <30min | 13,406 | 96.80% |
| <1hr | 404 | 2.92% |
| <2hr | 39 | 0.28% |
| Total | **13,849** |  |

n= All Patrons tested from July 17 2021 to Jan 31, 2022.
N.B. After October 4, only patrons with either positive, cancelled or invalid Cue results had an additional swab collected for lab-based RT-PCR.

## Discussion

This study sought to ascertain the real world efficacy of a screening program using the Cue POCT compared with RT-PCR in predominantly asymptomatic individuals. No false negatives were noted in this study cohort suggesting that the highly sensitive Cue POCT can rapidly and accurately detect cases. These performance characteristics have significant utility within a screening program for any large group gatherings (i.e workplace, congregate events, residences such as long term care homes, dormitories, etc) by rapidly identifying asymptomatic cases and promoting informed isolation and cohorting procedures which reduce the likelihood of transmission, and promote safe gatherings and business continuity. Additionally, the specificity of Cue POCT indicates that false positives are infrequent (high positive predictive value), decreasing the risk of unnecessary quarantine and the need for lab-based RT-PCR confirmatory testing which can inherently delay social gatherings **(12)**.

Early and accurate detection, particularly with Omicron, and more recent variants has been challenging due to the poor sensitivity of antigen testing as reviewed in multiple commentaries **(13,14,15)**. These observations are corroborated by findings described in **Adamson et al**, 2022 **(16)** as discordant antigen test results were frequently noted in both symptomatic and asymptomatic individuals when compared to PCR and molecular testing. The sensitivity of the Cue POCT indicates it is an accurate proxy for traditional lab-based PCR testing with the added advantage of rapid results. The performance of Cue POCT revealed accurate detection of early infection (lower viral load) which reduces the need for frequent testing, as recommended for antigen-based screens **(13,17,18)**. Together, these features position the Cue POCT as a valuable tool for asymptomatic screening with a superior value proposition for accuracy, cost, resources and time compared to antigen testing. The findings of this study also confirm that the real world clinical sensitivity of the Cue COVID-19 POC tests for SARS CoV2 wildtype and variants was much closer to gold standard RT-PCR testing than antigen testing.

The findings of this study can further be extrapolated to considerations of “Test to Treat” initiatives (19). As the pandemic has evolved, novel treatment options have become available, including Molnupiravir and Paxlovid, as well as monoclonals. A new feature of Cue allows the user to directly connect to a platform for treatment, interface with a health care provider, receive a prescription, and have it delivered directly to their residence (20). The inclusion of Cue POCT in Test to Treat initiatives holds the potential for synergistic benefits by improving time to diagnosis, accelerating clinical management, minimizing delays in treatment initiation and ultimately promoting treatment effectiveness with rapid viral clearance, reduced complications and lower hospitalization rates. Narrowing the time interval from test to treatment accelerates how quickly the patient can begin convalescence and return to work, which reduces overall economic stress, and has particular benefit to at-risk communities where other health and community resources, including testing, can be limited.

The cost of molecular tests is higher than the relative cost of antigen tests. However, when factoring in the performance of the tests, particularly in asymptomatic individuals, and the frequent need to repeat antigen testing to have similar detection of positive cases, the cost of molecular testing is comparable to antigen testing, and less disruptive to work flow. Additionally, when compared to lab-based PCR testing, POC molecular testing, as deployed in this study, Cue POCT did not incur many of the logistical costs seen in RT-PCR testing. Delays in lab-based PCR results can also compromise early initiation of treatment and associated health outcome benefits.

Potential limitations of the current study include the use of health care workers for Cue specimen collection, in contrast to self-collection of nasal specimens by individuals without formal professional training. Cue POCT has EUA authorization for “over the counter” sale in the US whereas Health Canada approval during the study period limited specimen collection and testing by health professionals only. In contrast, there may be a learning curve for untrained individuals to perform reliable self-swabbing which could impact downstream test performance. However, Cue provides an in-app video, step-by-step visual mobile app instructions and printed instruction sheets to ensure appropriate sampling methods, and there is ample data with other systems to show self-swabbing is reliable, safe, and comparable to health care worker collection. Other limitations of the current study include the lack of direct comparison to antigen testing, lack of viral culture and sequencing data to identify specific variants, and the lack of direct economic analysis. However, the study does represent one of the largest sets of data comparing lab-based RT-PCR to point of care molecular testing in a real world population of largely asymptomatic individuals.

The results of the current study underscore the success of Cue POCT in a community setting and the potential for broader population benefits, particularly in congregate settings and in high-risk populations. Early case identification, prior to symptom onset, has the potential to curtail transmission and limit the immense burden of disease complications, healthcare costs and socio-economic disruptions. The accuracy of Cue POCT results and the ease of use of the digitally-enabled Cue POCT device and mobile Cue Health app testing provide a cost-effective alternative to lab-based PCR testing and serial antigen testing. Additionally, the high sensitivity and specificity provide actionable results with confidence to employers and businesses seeking to proactively identify cases and prevent further spread of COVID. The point of care testing platform assessed in the current study has new capabilities to coordinate treatment through the mobile application, which can have a dramatic impact on time to treatment and how to manage the on-going pandemic.

## Data Availability

All data produced in the present study are available upon reasonable request to the authors

